# Modelling exposure to aerosols from showers: implications for microbial risk assessment

**DOI:** 10.1101/2025.01.30.25321398

**Authors:** Lizhan Tang, Antonia Eichelberg, Franziska Böni, Kerry A. Hamilton, Émile Sylvestre, Frederik Hammes, Timothy R Julian

**Affiliations:** Eawag, Swiss Federal Institute of Aquatic Science and Technology, 8600 Dulbendorf, Switzerland; Department of Environmental Systems Sciences, ETH Zulrich, 8092 Zürich, Switzerland; School of Sustainable Engineering and the Built Environment, Arizona State University, Tempe, Arizona, USA; Biodesign Institute Center for Environmental Health Engineering, Arizona State University, Tempe, Arizona, USA; Swiss Tropical and Public Health Institute, 4123 Allschwil, Switzerland; University of Basel, 4055 Basel, Switzerland

## Abstract

Inhalation of aerosols produced during showering exposes people to chemical and microbial contaminants present in the water. To improve quantitative estimates of exposure and to inform the efficacy of potential interventions to reduce exposures, we conducted empirical measurements of aerosol concentration and size distribution during showering events. We measured aerosol count concentrations and size distributions with an aerodynamic particle sizer over the duration of mock showering events under various conditions, including different water temperatures and different showerhead types (conventional and rain showers). The empirical data were then used to fit a mass balance model to obtain aerosol generation rates and decay rates for each aerosol size class through least square fitting. We observed an initial high peak concentration of aerosols under hot water conditions relative to cold water conditions which resulted in a rapid increase in aerosol exposure during the first 1-2 minutes of showering. This suggests that people showering in hot water conditions will have a potentially increased exposure during the first 1-2 minutes. The model-fitted values suggest large inter-experiment variation in estimated aerosol generation and decay rates, even among triplicates of the same showering conditions. Current exposure assessment approaches assume constant aerosol concentrations during showers which might lead to miscalculated cumulative risk. Thus, considering aerosol dynamics is beneficial during shower exposure assessments to inform risk management interventions. The data set and associated modeling results provided can support this, as they can be readily integrated into microbial risk assessment for waterborne pathogens such as *Legionella* spp., nontuberculous *Mycobacteria* (NTM) and *Pseudomonas aeruginosa*.

## 1. Introduction

Waterborne pathogens in drinking water systems such as *Legionella* spp., *Mycobacterium* spp. and *Pseudomonas aeruginosa* increase risks for respiratory diseases during daily activities, such as showering and toilet flushing (Dean & Mitchell, 2020; Hamilton et al., 2017). Inhalation of contaminated aerosols is the major route of exposure, with showers perceived to be of particular concern due to the generation of, and resultant exposure to, high concentrations of inhalable aerosols (Falkinham III, 2020; Hamilton et al., 2019; Shen et al., 2022). Understanding fate and transport of aerosols in showers can provide insights into exposure pathways leading to respiratory diseases and contribute to the design of effective interventions.

Estimates of aerosol concentrations, fate, and transport in showers are driven by data collected empirically. Parameters related to aerosol concentrations were usually identified as one of the most sensitive parameters for risk assessment (Tang et al., 2024). Empirical studies have quantified shower aerosol generation rates and identified factors that influence the aerosol concentration and distribution profile such as water flow rate, water temperature, water pressure, humidity, showerhead characteristics, and ventilation conditions (Cowen & Ollison, 2006; Estrada-Perez et al., 2018; Niculita-Hirzel et al., 2021; O’Toole et al., 2009; Xu & Weisel, 2003; Zhou et al., 2019). Some studies, primarily focused on chemical exposure assessment, collected data on aerosols smaller than 2.5 μm, a size class particularly relevant due to its deposition in the alveoli and bronchi (Xu & Weisel, 2003; Zhou et al., 2007). For aerosols larger than 2.5 μm, many current studies applied general aerosol generation rates or partitioning coefficients (ratio of pathogen in air relative to water) for all respirable size classes. (Cowen & Ollison, 2006; Estrada-Perez et al., 2018; Niculita-Hirzel et al., 2021; Zhou et al., 2019). Aerosols ranging between 1 to 10 μm are normally large enough to encapsulate microorganisms and small enough to deposit at alveoli. Moreover, aerosols larger than 5 μm are significantly less likely to deposit at alveoli compared to smaller aerosols(Heyder et al., 1975; Heyder et al., 1973; Hussain et al., 2011). Therefore, size-resolved generation rates for aerosols within this range are needed for reliable microbial risk assessments.

Microbial risk assessments rely on aerosol concentration modeling approaches including volumetric estimation or partitioning coefficient approaches to model exposure to respiratory pathogens for shower scenarios (Hamilton et al., 2017; Hamilton et al., 2019; Sales-Ortells & Medema, 2014). Most studies applying these approaches typically measure either aerosols or microorganisms as data input (Estrada-Perez et al., 2018; Hines et al., 2014; Zhou et al., 2007). Comparatively, few studies investigate both aerosol size distribution and microorganisms in air using impactor cultures for shower-like systems (Allegra et al., 2016; Allegra et al., 2020). These approaches typically assume constant aerosol concentration over the entire shower duration. A consequence of constant aerosol concentration estimation is the potential to overestimate or underestimate risk, because assumed constant aerosol concentration corresponds to steady state, while aerosol concentration at the growth state (period when aerosol concentration is continuously increasing) is usually ignored. An alternative to volumetric estimation and partitioning coefficient approaches is mass balance model approach, which can incorporate multiple aerosol transport process such as deposition and ventilation following the conservation of mass principle, and therefore be more generalizable. Previous studies focusing on chemical exposure assessment have shown that one compartment models can properly describe dynamic changes of aerosol concentrations (Cowen & Ollison, 2006; Zhou et al., 2007). Through fitting the mass balance model to experimental measured aerosol data, aerosol generation rates ranging from 4 to 2841 μg/m^3^/min were obtained depending on different aerosol size classes and showerhead characteristics (Cowen & Ollison, 2006; Xu & Weisel, 2003; Zhou et al., 2007). Compared to a conventional volumetric estimation approach, a mass balance model could be used to study the impact of aerosol dynamics on risk assessments and better inform risk management strategies through incorporation of different aerosol removal mechanism.

In this study, we advanced empirical data collection and modeling of aerosols in showers. Specifically, we (1) investigated the temporal variation of aerosol concentration and size distribution of aerosols under selected conditions with shower experiments, 2) estimated size-resolved parameters for aerosol generation rates that can be directly used in exposure and risk assessment models, and (3) developed and applied a mass balance model that describes temporal variations of aerosol concentrations and corresponding exposures for people showering.

## 2. Methods

### 2.1 Experimental overview

Empirical measurements of aerosol concentration and size distribution were collected in the bathroom of a fitness center in Dübendorf, Switzerland. The shower stall has a dimension of 0.9m (length) x 1.2m (width) x 2.3m (height). The shower stall was closed with a glass door, and air exchange was achieved by a ventilation system installed at the top of the shower stall. A conventional showerhead and a rain showerhead (Arwa, Switzerland) made of stainless steel were installed in the shower stall. Both types of showerheads have a water pressure of 3 kPa and a nozzle diameter of 1mm. The conventional showerhead has 65 nozzles and rain showerhead has 210 nozzles. The rain showerhead was set at the ceiling, 2.3m above ground. The conventional showerhead was also fixed at the position of the rain showerhead to ensure aerosols were emitted at the same position. A 1.7m mannequin was positioned right below the rain and conventional showerhead to simulate the presence of a person during the shower and to account for secondary formation of aerosols due to water splashing onto human skin.

Two temperatures (moderately cold at 24-26 ℃ and moderately hot at 39-41℃) and two showerhead types (conventional and rain) were tested for aerosol concentration and size distribution. The maximum water flow rates were 10L/min for the rain showerhead and 8L/min for the conventional showerhead. Three positions (1m above ground 1.8m above ground and 2.2m above ground) were selected as sampling points for aerosol size distribution and concentration measurements (aerodynamic particle sizer APS 3321, TSI, Minnesota). Aerosols were sampled using tubes connected to the inlet port of APS with a total flow rate of 5L/min (1L/min for aerosol sampling and 4L/min for sheath air). The outputs of APS are size distribution normalized by aerosol size and aerosol count for each aerosol size class, which included 51 intervals within the size range of 0.5 to 20 μm.

Measurements were collected for 5 minutes prior to the shower being turned on to measure the background aerosol concentration. Then, the shower was turned on for 10 minutes to mimic a typical shower length. Then, the shower was turned off but aerosols were still measured for an additional 5 minutes to allow measurements of the decay of aerosol concentrations back to the background. To measure the ventilation rate, carbon dioxide (CO_2_) was used as a tracer gas (Batterman, 2017). Carbon dioxide was injected from a compressed gas cylinder into the shower stall until an initial concentration of 3000-4000 ppm was reached. This concentration was well below the American Conference of Governmental and Industrial Hygienists’ recommended 8-hour time weighted average threshold Limit Value (Association, 1978) while also being sufficiently above background to enable estimation of ventilation rate. Before measuring CO_2_ concentration, a fan was used to ensure the well-mixed air. Decay of CO_2_ concentration was then recorded using Vitales 501 CO_2_ monitor (Vitales, Switzerland). Through fitting an exponential decay model to the measured concentrations over time, the ventilation rate was estimated (Zhou et al., 2007). The carbon dioxide monitor also recorded relative humidity and temperature every 20 seconds.

### 2.2 Aerosol concentration modeling

#### 2.2.1 Mass balance model

To estimate exposure for a person enclosed within a shower stall, a mass balance model was used (Zhang et al., 2020). Assuming a closed system following the principle of mass conservation and accounting for the generation rate from the shower and decay from ventilation, deposition, and other processes, the mass concentration of aerosol was predicted as follows:

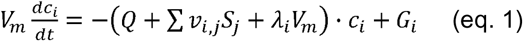

where *V_m_* is the volume of the shower stall (m^3^), *c_i_* is the mass concentration of aerosol of size i (μg/m^3^), *t* is the exposure duration (min), *Q* is the ventilation rate or air flow rate (m^3^/min); *S_j_* is the area of vertical, upward-facing, and downward-facing surface j within the shower stall (m^2^); *v_i,j_* is the deposition velocity of aerosol of aerosol of size i for the surface j (m/min); *λ_i_* is the aerosol residual decay rate caused by other removal process excluding deposition and ventilation rate for aerosol of size i (1/min); *G* is the generation rate of aerosol of size i (μg/min). The deposition velocities were calculated following the method proposed by (Lai & Nazaroff, 2000).

#### 2.2.2 Model calibration

The mass balance model was fitted to measured aerosol concentrations for each aerosol size class using the least squares method (Marcoline et al., 2020). Aerosol generation rate and aerosol decay rate were treated as unknown parameters during the model fitting. The fitting process was conducted using the differential equations solver Berkeley Madonna (University of California at Berkeley, 9.1.19) with the Rosenbrock algorithm for stiff differential equations.

#### 2.2.3 Estimating exposure to respirable aerosols

The potential inhalation dose of aerosols for a single shower event was calculated using the following equation:

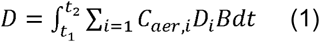

where *D* is the inhalation dose of aerosols (g), *C_aer,i_* is the mass concentration of aerosols of size i (g/m^3^), *B* is the inhalation rate (m^3^/min), and *D_i_* is the deposition efficiency of an aerosol of size i at human alveoli, *t_1_* is the time when people enter shower stall and *t_2_* is the time when people leave shower stall (min).

Exposure modeling, including estimating uncertainty and variability of model output, was conducted using Monte Carlo simulations in R v.4.0.2. (www.rproject.org), based on 10,000 iterations.

## 3. Results

### 3.1 Aerosol size distribution

Aerosol size distributions were generally consistent throughout the shower. We observed no significant change in the geometric mean diameter of aerosols generated from the start of the shower (1^st^ minute of sampling) and end of the shower (10^th^ minute of sampling) (Table 1) (p=0.02, t-test), but differences were observed when comparing the geometric mean diameters between the cold-water scenarios and hot water scenarios (p=1.7×10^−9^, t-test). Geometric mean diameters for aerosols generated using the conventional showerhead at the 1^st^ and 10^th^ minutes were similar to the geometric mean diameters generated using the rain showerhead (*p* = 0.71, t-test). For cold water scenarios, the majority of aerosols generated by the two types of showerheads were smaller than 5 μm and smaller aerosols accounted for a higher fraction of total number concentrations (Figure 1). Comparatively, aerosols generated by hot water showers were larger than aerosols generated by cold water showers and were characterized by a bimodal distribution (Figure 1 and Table 1).

**Table 1.** Summary of aerosol size distribution under selected conditions. t=1^st^ minute represents the beginning of the shower after the first five minutes of measuring background concentrations and t=10^th^ minute represents the end of the shower before measuring aerosol decay.

| Experiment run | Water temperature (°C ) | Showerhead type | Water flow rate ( L/min ) | Geometric mean diameter of generated aerosols (µm) |  |
| --- | --- | --- | --- | --- | --- |
|  |  |  |  | Time=1<sup>st</sup> minute | Time=10<sup>th</sup> minute |
| 1 | 25.9 | Rain | 10 | 0.95 | 1.16 |
| 2 | 25.8 | Rain | 10 | 0.96 | 0.96 |
| 3 | 27.3 | Rain | 10 | 0.97 | 1.02 |
| 4 | 29.3 | Conventional | 8 | 0.85 | 0.96 |
| 5 | 27.8 | Conventional - | 8 | 1.12 | 0.94 |
| 6 | 26.6 | Conventional - | 8 | 0.97 | 1.07 |
| 7 | 39.7 | Rain - | 10 | 5.03 | 6.12 |
| 8 | 41.9 | Rain - | 10 | 5.29 | 6.48 |
| 9 | 41.0 | Rain - | 10 | 5.22 | 5.03 |
| 10 | 40.0 | Conventional - | 8 | 4.10 | 4.81 |
| 11 | 40.5 | Conventional - | 8 | 5.51 | 6.36 |
| 12 | 39.5 | Conventional - | 8 | 3.96 | 4.38 |

**Figure 1.**
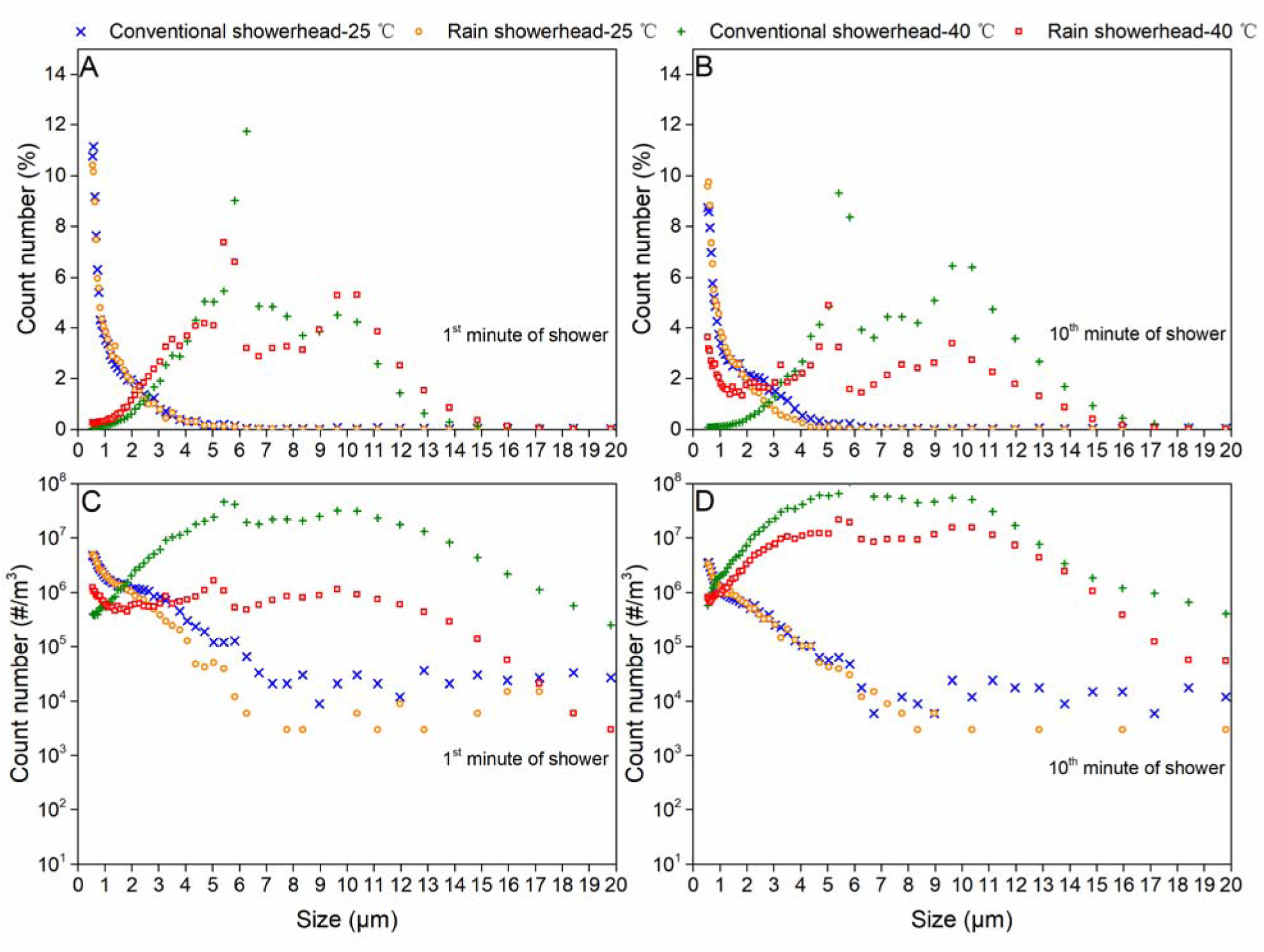
Size distribution as a percent of the total number (A) and (B), as well as total number concentration (C) and (D). The figure shows aerosol size data for one of the triplicates at each condition sampled in a shower stall with mannequin present.

### 3.2 Concentration of aerosols over time

The concentration profile of aerosols generated during and after the shower can be divided into 3 phases: 1) growth state (1-2 minutes after the shower was turned on), 2) steady state (1-2 minutes after shower is turned on), and 3) decay state (after the shower was turned off). For cold water scenarios, the concentration of aerosols increased exponentially during the transition state (Figures 2,3). In many scenarios, a plateau in aerosol concentrations indicating steady state was reached. In other scenarios, the aerosol concentrations continuously increased until the end of the shower and a steady state concentration was not reached (Figure S1, Figure S2). The specific scenario depended on the air exchange rate within the shower, with higher air exchange rates reaching steady state faster than lower air exchange rates, as expected from the mass balance model.

**Figure 2.**
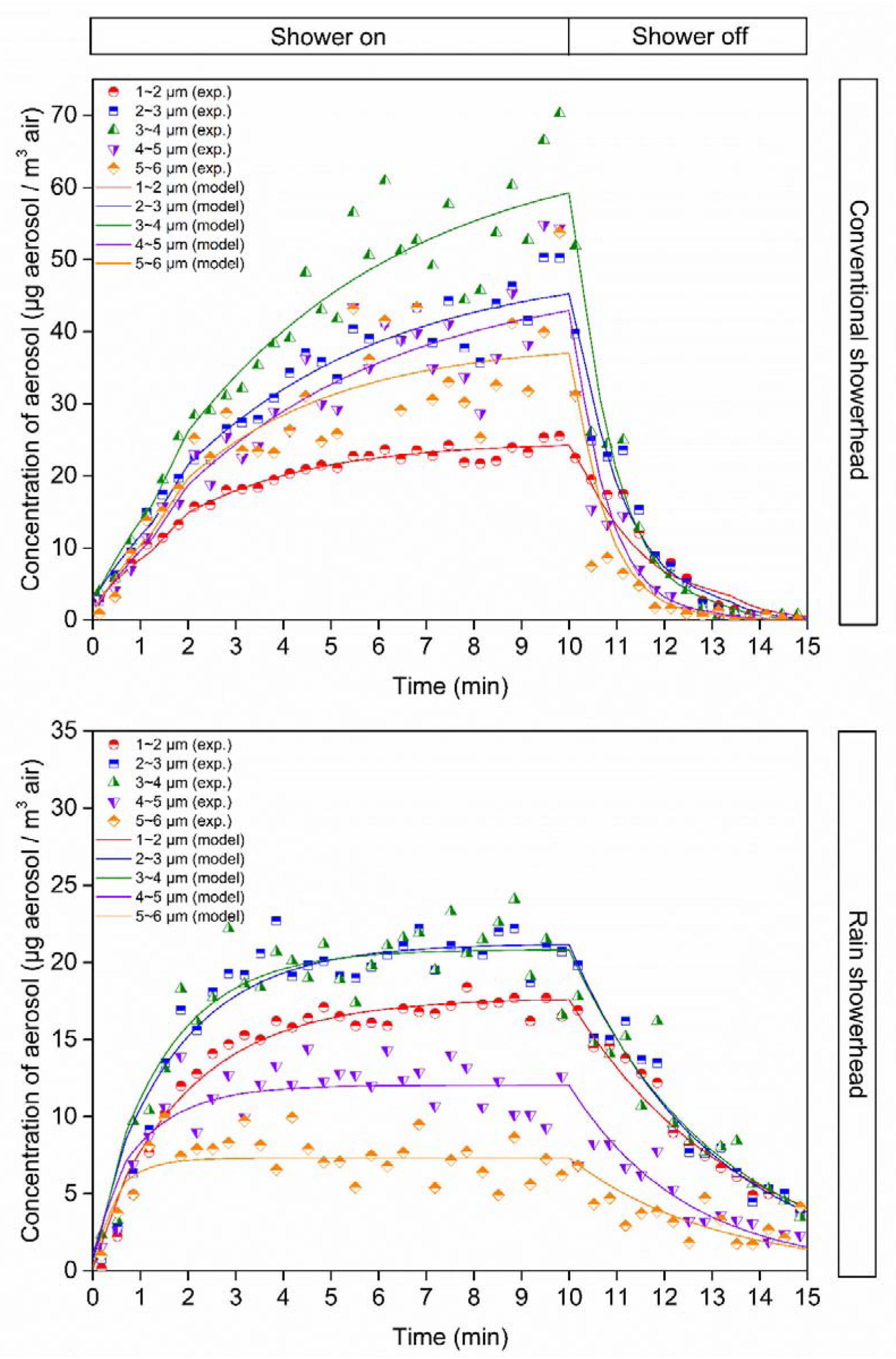
Concentration of aerosols of each size class for one of the triplicates at cold water temperature. The term exp refers to the experimentally collected data, and model refers to the model fit values.

**Figure 3.**
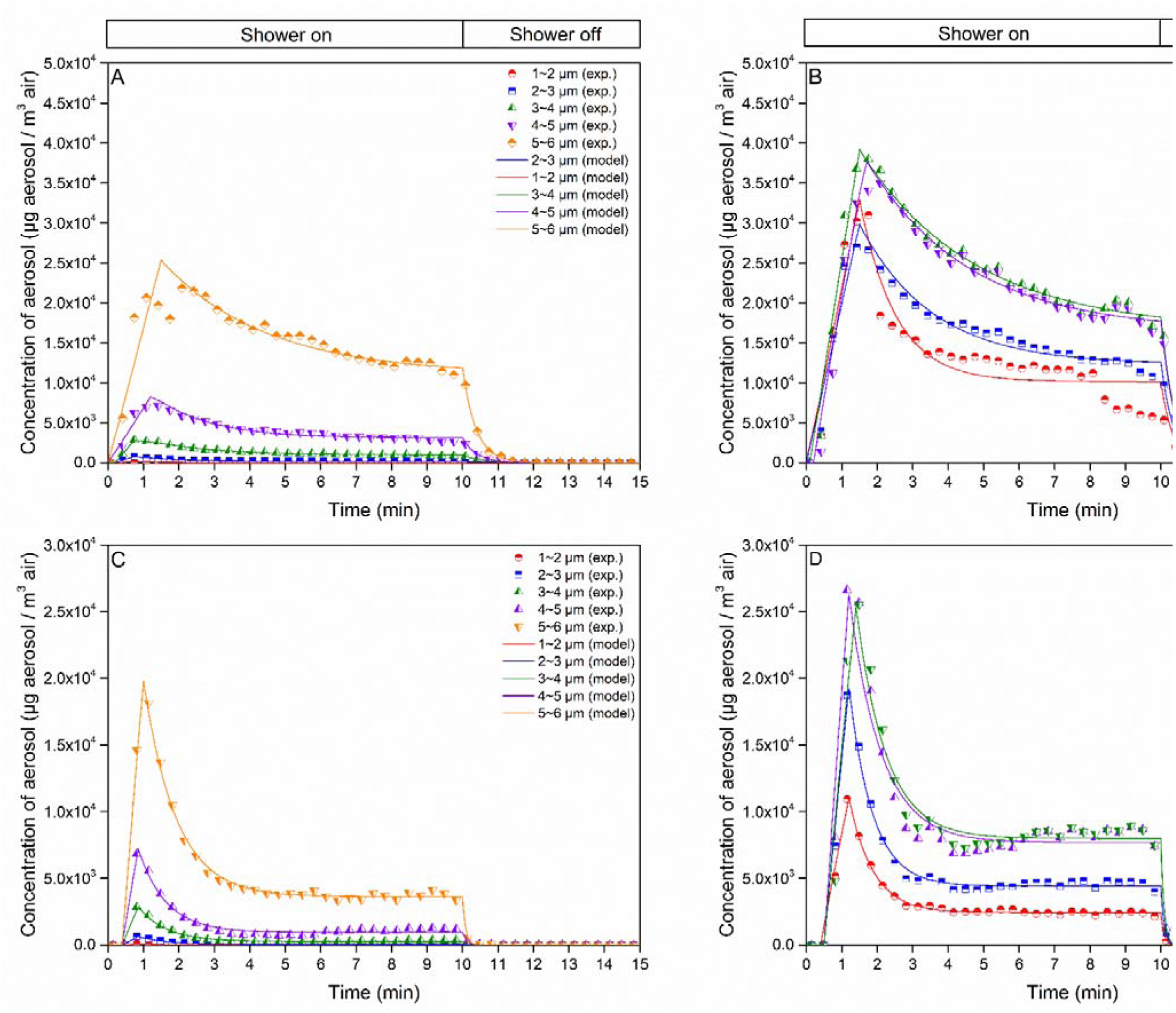
Concentration of aerosols for one of the triplicates at hot water temperature. The term exp refers to the experimentally collected data, and model refers to the model fit values.

For hot water scenarios, an almost instantaneous increase of aerosol concentration after the shower was turned on was observed in all experiments (Figure 3). After approximately 1 minute, a peak concentration was reached and then aerosol concentrations decreased exponentially until reaching a steady state (Figure 3). Notably, the peak concentration was sometimes 2-3 times higher than the steady state concentration (Figure 3). After the shower was turned on, both air temperature and relative humidity (RH) also gradually increased (Figure S5. For hot water scenarios, the highest air temperature at steady state was around 30L and the relative humidity gradually increased to 95% (Figure S5). For cold water scenarios, the rapid increase in aerosol concentrations was not observed, and the air temperature eventually reached a water temperature of approximately 25℃. The background RH, ranging from 50% to 80%, gradually increased to 85% under cold water conditions and 95% under hot water conditions (Figure S5).

### 3.3 Aerosol generation rates and decay rates

A mass balance model was fit to measured aerosol concentrations to get size-resolved aerosol generation rates and decay rates (Figure 2, Figure 3). The model fitting was conducted on each of the three phases separately to allow estimation of distinct aerosol generation rates and decay rates. For cold water scenarios, we assumed a constant aerosol generation rate for the growth and steady state phases (i.e. a single generation rate for the entire shower). For hot water scenarios, because of the empirically observed rapid initial increase of aerosol concentration, we modeled the growth and steady states as two distinct aerosol generation rates (i.e. a higher generation rate for the first 1-2 minutes and a lower generation rate after the first 1-2 minutes). For both cold and hot water scenarios, we applied two distinct aerosol decay rates: one rate for when the shower was turned on and a separate decay rate for when the shower was turned off, considering environmental conditions such as ambient temperature and relative humidity can be different when the shower is on or off.

Total aerosol generation rates for hot water scenarios were up to three orders of magnitude higher than those for cold water scenarios (Table 2, Table 3). In hot scenarios, aerosols ranging from 5-6 μm have the highest generation rates both by count and mass. Notably, in hot water conditions, the aerosol generation rates during the transition state were 2-4 times higher than those during the steady state.

**Table 2.** Model-fitted aerosol generation rates and decay rates under cold water conditions. Aerosol decay rate excludes ventilation rate and deposition rate. SD refers to Standard Deviation, Rain refers to the rain showerhead type, and Conventional refers to the conventional showerhead type.

| Showerhead<br>type | Flow rate<br>( L/min ) | Size range<br>( μm ) | Aerosol generation<br>rate ( μg/min ) |  | Decay rate ( 1/min ) |  |  |  | Water temperature<br>(°C ) |  |
| --- | --- | --- | --- | --- | --- | --- | --- | --- | --- | --- |
|  |  |  |  |  | Growth state and<br>steady state |  | Decay stage |  |  |  |
|  |  |  | Mean | SD | Mean | SD | Mean | SD | Mean | SD |
| Rain | 10 | 1-2 | 19.62 | 2.69 | -0.01 | 0.15 | 0.11 | 0.17 | 26.3 | 0.8 |
| Rain | 10 | 2-3 | 25.88 | 3.97 | -0.03 | 0.15 | 0.15 | 0.16 | 26.3 | 0.8 |
| Rain | 10 | 3-4 | 29.07 | 6.02 | 0.00 | 0.19 | 0.12 | 0.17 | 26.3 | 0.8 |
| Rain | 10 | 4-5 | 20.32 | 6.00 | 0.08 | 0.25 | 0.16 | 0.22 | 26.3 | 0.8 |
| Rain | 10 | 5-6 | 18.42 | 11.95 | 0.45 | 0.63 | 0.12 | 0.17 | 26.3 | 0.8 |
| Conventional | 8 | 1-2 | 21.32 | 1.57 | 0.06 | 0.09 | 0.19 | 0.22 | 27.9 | 1.4 |
| Conventional | 8 | 2-3 | 33.25 | 9.42 | -0.03 | 0.06 | 0.35 | 0.33 | 27.9 | 1.4 |
| Conventional | 8 | 3-4 | 50.98 | 31.06 | 0.01 | 0.03 | 0.45 | 0.40 | 27.9 | 1.4 |
| Conventional | 8 | 4-5 | 39.49 | 24.98 | -0.08 | 0.16 | 0.54 | 0.50 | 27.9 | 1.4 |
| Conventional | 8 | 5-6 | 56.97 | 55.91 | -0.04 | 0.63 | 0.60 | 0.48 | 27.9 | 1.4 |

**Table 3.** Model fitted aerosol generation rates and decay rates under hot water conditions. Aerosol decay rate excludes ventilation rate and deposition rate. SD refers to Standard Deviation, Rain refers to the rain showerhead type, and Conventional refers to the conventional showerhead type.

| Showerhead type | Size range (μm) | Aerosol generation rate ( μg/min ) |  |  |  | Decay rate ( 1/min ) |  |  |  | Water temperature |  | Flow rate ( L/min ) |
| --- | --- | --- | --- | --- | --- | --- | --- | --- | --- | --- | --- | --- |
|  |  | Growth state |  | Steady state |  | Steady state |  | Decay stage |  | (°C ) |  |  |
|  |  | Mean | SD | Mean | SD | Mean | SD | Mean | SD | Mean | SD |  |
| Rain | 1-2 | 6.9x10<sup>2</sup> | 5.9x10<sup>2</sup> | 2.6x10<sup>2</sup> | 2.9x10<sup>2</sup> | 2.6 | 2.3 | 8.0x10<sup>-1</sup> | 5.1x10<sup>-1</sup> | 40.9 | 1.1 | 10 |
| Rain | 2-3 | 4.5x10<sup>3</sup> | 4.1x10<sup>3</sup> | 2.1x10<sup>3</sup> | 2.3 x10<sup>3</sup> | 3.6 | 4.1 | 1.4 | 1.8x10<sup>-1</sup> | 40.9 | 1.1 | 10 |
| Rain | 3-4 | 2.3x10<sup>4</sup> | 2.4x10<sup>4</sup> | 9.7E+03 | 1.1x10<sup>4</sup> | 3.0 | 3.1 | 2.6 | 1.2 | 40.9 | 1.1 | 10 |
| Rain | 4-5 | 6.2x10<sup>4</sup> | 6.6x10<sup>4</sup> | 1.8x10<sup>4</sup> | 1.7x10<sup>4</sup> | 2.5 | 2.5 | 3.8 | 2.8 | 40.9 | 1.1 | 10 |
| Rain | 5-6 | 1.4x10<sup>5</sup> | 1.6x10<sup>5</sup> | 7.7x10<sup>4</sup> | 1.0x10<sup>5</sup> | 2.1 | 1.2 | 5.3 | 6.3 | 40.9 | 1.1 | 10 |
| Rain | 6-7 | 7.4x10<sup>4</sup> | 8.8x10<sup>4</sup> | 1.4x10<sup>4</sup> | 4.9x10<sup>3</sup> | 1.2 | 1.1 | 5.6 | 6.6 | 40.9 | 1.1 | 10 |
| Rain | 7-8 | 8.2 x10<sup>4</sup> | 6.8 x10<sup>4</sup> | 3.1 x10<sup>4</sup> | 1.7 x10<sup>4</sup> | 1.3 | 8.7x10<sup>-1</sup> | 5.4 | 6.4 | 40.9 | 1.1 | 10 |
| Rain | 8-9 | 1.0x10<sup>5</sup> | 7.2x10<sup>4</sup> | 5.3x10<sup>4</sup> | 3.1x10<sup>4</sup> | 1.3 | 1.1 | 5.3 | 6.3 | 40.9 | 1.1 | 10 |
| Rain | 9-10 | 9.2x10<sup>4</sup> | 6.9x10<sup>4</sup> | 4.2x10<sup>4</sup> | 1.6 x10<sup>4</sup> | 1.2 | 9.1x10<sup>-1</sup> | 5.6 | 6.7 | 40.9 | 1.1 | 10 |
| Conventional | 1-2 | 6.1x10<sup>2</sup> | 4.0x10<sup>2</sup> | 2.3x10<sup>2</sup> | 2.9x10<sup>2</sup> | 9.6x10<sup>-1</sup> | 4.0x10<sup>-1</sup> | 1.5 | 4.2x10<sup>-1</sup> | 40.0 | 0.5 | 8 |
| Conventional | 2-3 | 3.5x10<sup>3</sup> | 3.2x10<sup>3</sup> | 9.3x10<sup>2</sup> | 1.2x10<sup>3</sup> | 6.8x10<sup>-1</sup> | 2.1x10<sup>-1</sup> | 1.8 | 8.7x10<sup>-1</sup> | 40.0 | 0.5 | 8 |
| Conventional | 3-4 | 1.5x10<sup>4</sup> | 1.5x10<sup>4</sup> | 3.6x10<sup>3</sup> | 5.1x10<sup>3</sup> | 4.8x10<sup>-1</sup> | 1.3x10<sup>-1</sup> | 2.3 | 7.1x10<sup>-1</sup> | 40.0 | 0.5 | 8 |
| Conventional | 4-5 | 3.2x10<sup>4</sup> | 4.0x10<sup>4</sup> | 7.5x10<sup>3</sup> | 9.1x10<sup>3</sup> | 4.8x10<sup>-1</sup> | 1.8x10<sup>-2</sup> | 2.5 | 7.1x10<sup>-1</sup> | 40.0 | 0.5 | 8 |
| Conventional | 5-6 | 8.5x10<sup>4</sup> | 1.1x10<sup>5</sup> | 2.8x10<sup>4</sup> | 4.1x10<sup>4</sup> | 4.7x10<sup>-1</sup> | 4.1x10<sup>-1</sup> | 2.3 | 8.9x10<sup>-1</sup> | 40.0 | 0.5 | 8 |
| Conventional | 6-7 | 9.0x10<sup>4</sup> | 1.1x10<sup>5</sup> | 1.8x10<sup>4</sup> | 1.6x10<sup>4</sup> | 5.7x10<sup>-1</sup> | 2.3x10<sup>-1</sup> | 2.3 | 1.2 | 40.0 | 0.5 | 8 |
| Conventional | 7-8 | 8.2x10<sup>4</sup> | 1.0x10<sup>5</sup> | 2.3x10<sup>4</sup> | 2.7x10<sup>4</sup> | 3.5x10<sup>-1</sup> | 3.1x10<sup>-1</sup> | 2.1 | 1.1 | 40.0 | 0.5 | 8 |
| Conventional | 8-9 | 7.6x10<sup>4</sup> | 7.7x10<sup>4</sup> | 2.8x10<sup>4</sup> | 3.8x10<sup>4</sup> | 2.4x10<sup>-1</sup> | 3.1x10<sup>-1</sup> | 2.0 | 1.1 | 40.0 | 0.5 | 8 |
| Conventional | 9-10 | 7.9x10<sup>4</sup> | 8.5x10<sup>4</sup> | 2.1x10<sup>4</sup> | 2.6x10<sup>4</sup> | 1.8x10<sup>-1</sup> | 2.2x10<sup>-1</sup> | 1.9 | 1.0 | 40.0 | 0.5 | 10 |

In the one compartment model, an additional parameter fit to the model was a parameter to estimate the residual aerosol decay rate (λ) to account for aerosol removal processes that occur in addition to the ventilation and deposition removal processes. This rate is intended to represent combinations of other potential removal processes, such as evaporation, condensation, coagulation, and thermophoresis. Residual aerosol decay rates close to 0 would indicate the aerosol removal processes beyond ventilation and deposition are negligible. Based on fitted values, the residual aerosol decay rates under cold water conditions (Table 2) were much closer to 0 than those under hot water conditions (Table 3). Specifically, we observed that fitted residual aerosol decay rates for the rain showerhead under hot water conditions were both higher and more variable than the other scenarios. We also observed higher residual aerosol decay rates during the decay phase (e.g., when the shower was turned off) compared to the transition and steady state phases (e.g., when the shower was turned on).

Aerosol generation rates were used to estimate aerosol exposures through inhalation under all tested conditions (Figure 4). The estimated cumulative doses for inhalation of aerosols under hot water conditions, calculated based on the area under the curve, was 2.3 mg for conventional showerhead and 3.5 mg for rain showerhead, which was two orders of magnitude higher than cold water estimates of 2.4×10^−2^ mg for conventional showerhead and 6.6×10^−3^ mg for rain showerhead. For hot water scenarios, the slope of increase for the initial 2 minutes was higher than the remaining period due to the initial peak concentration of aerosols during the transition phase (Figure 4). Comparatively, the slope of increase for cold water scenarios was much lower due to the low aerosol generation rates. After the shower was turned off, a continued exposure increase was observed for cold water scenarios while the exposure for hot water scenarios reached a plateau. This is because, under hot water scenarios, the aerosol concentration typically decreased dramatically, reaching almost undetectable once the showerhead is off, whereas the aerosol concentration decreased more slowly under cold water scenarios.

**Figure 4.**
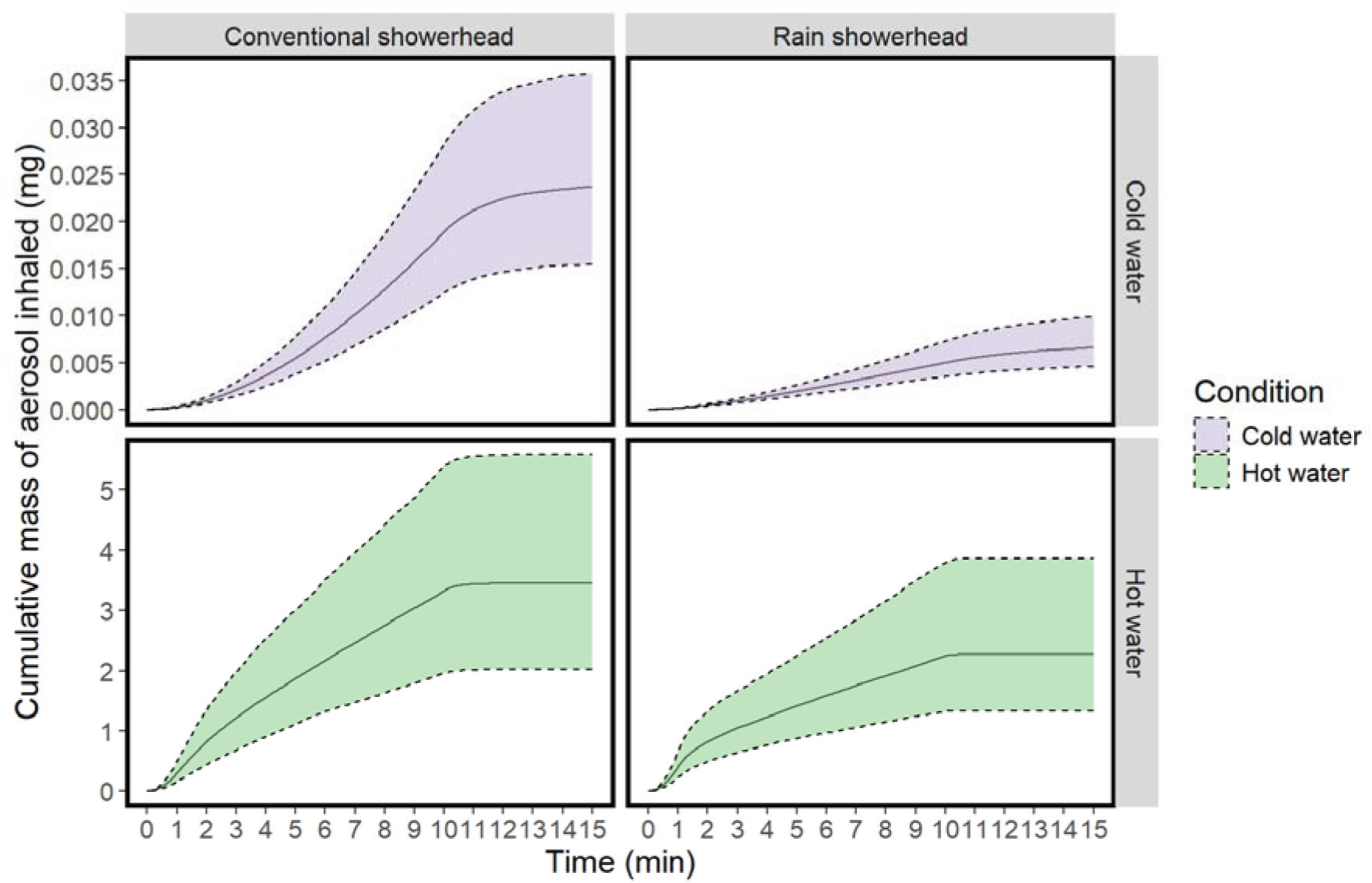
Estimated exposure in mass of aerosols from showers inhaled by a person over time. Solid line represents median value and dashed lines represent 5 percentile and 95 percentile values.

## 4. Discussion

### 4.1 Aerosol generation rates were more influenced by water temperature than by showerhead type

We observed in our study that the showerhead types had little influence on aerosol concentration and size distribution, whereas temperature had a larger effect. Our observation of a minimal impact of showerheads may be due to the study design, which looked at a single, constant flow rate for each shower type, and the flow rates for the two showerhead types were similar: 8 and 10 liters per minute. Studies have shown that showerhead characteristics such as water flow rate, water pressure, number of nozzles and spray angle can influence aerosol emission to different extents (Niculita-Hirzel et al., 2021; Zhou et al., 2007). However, the difference in aerosol generation rates caused by showerhead characteristics may be low, typically reported as within 1 log_10_ (Estrada-Perez et al., 2018; Pitell et al., 2024; Xu & Weisel, 2003). Comparatively, water temperature differences can result in 2-3 log_10_ differences in aerosol generations rates (Zhou et al., 2007). Our finding is consistent with previously reported results on the importance of temperature in aerosol generation rates, and highlights that risk mitigation strategies focused on reducing water temperatures may see greater reductions in aerosol exposures than strategies focused on changing showerhead characteristics. However, it should be noted that our study only focused on aerosol exposures without considering impact of testing conditions on microorganisms. Water temperature as well as water flow rate (e.g., water-saving showerheads) could also impact microbial densities and viability in aerosols which could also influence the final risk outcomes (Chattopadhyay et al., 2017; Niculita-Hirzel et al., 2022; Pitell et al., 2024).

The observed higher aerosol concentrations under hot water scenarios, particularly during the steady state phase, may be due to increased humidity. Higher water temperatures lead to faster evaporation and correspondingly increase the air humidity near the showerhead. The humidified hot air may carry small droplets to regions with lower temperatures and the temperature difference between droplet surface and surrounding air combined with high humidity may lead to condensation. Indeed, we found that under cold water scenarios, only droplets smaller than 6 μm were measured, which indicates that evaporation processes likely dominated relative to condensation.

### 4.2 Aerosol decay rates vary at different stages and different water temperature conditions

In our one compartment model, the non-zero values of fitted aerosol decay rates (λ) suggests that there are processes controlling aerosol removal beyond ventilation and deposition. Our finding is consistent with previous studies that also observed deviation of total aerosol decay rates from the sum of air exchange rate and deposition rate (Cowen & Ollison, 2006; Zhou et al., 2007). Additional aerosol removal process may include evaporation, condensation, and coagulation, as well as deposition caused by diffusion and thermal gradients (Hussein et al., 2005; Xu & Weisel, 2003). These processes can be impacted by the dynamic change of environmental conditions such as temperature and relative humidity and therefore lead to large variations in our fitted aerosol decay rates.

Our model fitted larger aerosol decay rates at the decay stage compared to steady state, suggesting that other process in addition to deposition and ventilation process also play a role on aerosol removal when the shower was turned off. Our finding contradicts previous studies which observed no significant difference between aerosol decay rates and the sum of air exchange rate and deposition rate after the shower was turned off (Cowen & Ollison, 2006; Xu & Weisel, 2003). A possible explanation for this difference is that air exchange rates and aerosol generation were monitored separately in previous studies. In our study, we monitored aerosol concentration and air exchange rate simultaneously for each shower experiment, allowing higher resolution in our estimate of the contributions of ventilation rate to aerosol decay processes. Higher aerosol decay rates during the declining stage are reasonable. The lower relative humidity after a shower is turned off can promote evaporation of aerosols, explaining the higher aerosol decay rates during the decay state.

Our modeled aerosol decay rates under hot water conditions were higher than those under cold water conditions, suggesting other processes beyond ventilation and deposition may dominant aerosol removal under hot water conditions. A possible explanation for faster decay of aerosol concentrations under hot water condition than cold water conditions is that thermal plumes can carry smaller aerosols (<2.5 μm) upward, increasing the concentrations near the sampler (and, therefore, the person in the shower). Once the shower is turned off, those aerosols cannot be captured by sampling equipment without the rising plume, and instead decrease suddenly as they settle faster (Zhou et al., 2007).

### 4.3 Implications of dynamic model of aerosol concentration on risk assessment

Our observed initial peak aerosol concentrations under hot water conditions suggest the potential for a high exposure to aerosols within the first few minutes of turning on a shower. Similar instantaneous increases of aerosol concentrations were also observed in previous studies (Xu & Weisel, 2003; Zhou et al., 2007). There are two possible explanations for the instantaneous increase of aerosol concentrations under hot water conditions. First, hot water in some systems may be under a higher pressure than cold water systems, which may lead to the observed pulse input of aerosols after the shower was turned on. A second possible explanation is that air near the showerhead is quickly heated by the hot water, and as such forms a temperature gradient between the emission source and sampling point. The generated aerosols may be pushed from higher temperature regions to lower temperature regions through thermophoresis (Dhanraj et al., 2019). A potential intervention to reduce exposure to the observed high peak is to avoid entering the shower until aerosol concentrations have stabilized.

Avoiding the first few minutes of aerosol generation may also have impacts on exposures to microorganisms. The concentration of waterborne pathogens such as *Legionella* spp. have been observed to be highest in the first draw sample during flushing, likely due to biofilm detachment and favorable growth conditions in the last meter of pipes connecting to the point of use (Grimard-Conea et al., 2022; Shen et al., 2017). A combination of factors including an elevated of first draw pathogen concentration with an initial peak concentration of aerosols may result in higher exposures early in the shower period and already lead to a risk above an acceptable threshold. This poses a potential opportunity to develop water and aerosol sampling strategies focusing on first draw concentration for routine monitoring program.

### 4,4 Limitations and further improvements for aerosol modelling

The study findings provide a foundation for incorporating dynamic aerosol generation rates in showers into chemical and microbial risk assessments, but we note a number of limitations of the data provided. First, the dynamics observed are linked to the physical characteristics of the shower studied: other shower stalls with different dimensions, shower head types, flow rates, and ventilation rates may produce different dynamics. We note that despite this limitation, our modeling approach provides generalizable findings consistent with previous publications as our experimental conditions and shower stall size are within the previously reported ranges which are typical to mimic daily showers (Cowen & Ollison, 2006; Xu & Weisel, 2003). Further, the modeling approach is easily extendable to quantify generation and decay rates from empirical data collected in the future, particularly if the conditions to be modeled are determined to be sufficiently different from the conditions studied here. Second, the study highlights that under some scenarios, incorporation of aerosol removal processes beyond ventilation and deposition may be important for understanding fate and transport of aerosols in showers. The one compartment model investigated decay from sources other than ventilation and deposition by inclusion of an additional residual decay rate. In some scenarios, this residual decay rate was not negligible, suggesting additional aerosol removal processes could be integrated into the mechanistic model. There are several models focusing on aerosol dynamics that also incorporate evaporation and condensation process to account for their evolution of size distribution (Dhawan & Biswas, 2021; Hussein et al., 2005; Seigneur et al., 1986). However, such evaporation process should be carefully considered: much larger initial aerosol size means pathogen enrichment might happen due to the shrinkage of aerosols. The relative impact of pathogen enrichment on microbial risk assessment is an area of potential future research including in both laboratory and modeling studies (Armstrong & Haas, 2008; Armstrong & Haas, 2007).

Third, the mass balance model used here is also limited in that it does not account for the shifts of aerosol size distribution over time as well as spatial variation of aerosol concentrations within shower stalls. Our results for aerosols sampled at different locations showed stratification of aerosols due to thermal plume which suggest that ideal well-mixed condition is not meet (Figure S6). Computational fluid dynamic modeling approaches are an alternative to one-compartment models that can track the behavior of each individual aerosol. Such aerosol dynamic modelling approaches focus on the behavior of individual aerosols by calculating particle velocity from an initial velocity field, air velocity and external forces such as gravity and friction. Combined with size change of individual aerosols through an evaporation model, the spatial position and size over time of each individual particle can be estimated (Dhawan & Biswas, 2021). Alternatively, the initial particle velocity field can be measured through particle tracking velocimetry (PTV) (Estrada-Perez et al., 2018) as parameter input for the aerosol evolution model. In addition, CFD models are more generalizable than mass balance models as they require less scenario-specific data inputs which is a common limitation in applying current risk assessment frameworks.

## 5. Conclusions

This study integrates empirical data collection and modeling on aerosol size distributions from showers under multiple scenarios, including hot and cold water for both conventional and rain showerheads. The resulting data set and fitted model parameters can help to inform microbial and chemical exposure assessments. Notably, the study highlighted that water temperature has a larger influence on exposures than showerhead type studied here. Under hot water conditions, large aerosol generation rates occurring in the first minute of a shower indicate a potential high exposure risk during the first-minute of flushing. Model fitting highlighted that although aerosol decay rates are driven primarily by ventilation and aerosol deposition, in some scenarios other removal processes such as deposition by thermal gradients likely influence the concentration and mass of aerosols inhaled during showering. The findings provide meaningful data for further risk assessment frameworks and help inform sampling strategies for routine monitoring program.

## Data Availability

The data produced in the present study are available upon reasonable request to the authors.

## Acknowledgements

We thank Jing Wang for aid in the experimental design and analysis.

## 7. Author Contributions

The author contributions below are according to the CRediT statement. LT: Conceptualization, Methodology, Software, Validation, Formal analysis, Investigation, Data Curation, Visualization, Writing-Original Draft, Writing - Review & Editing. AE: Methodology, Investigation, Writing – Review and Editing. FB: Methodology, Investigation, Writing – Review and Editing. KH: Conceptualization, Methodology, Writing – Review and Editing. ES: Conceptualization, Methodology, Writing – Review and Editing. FH: Conceptualization, Methodology, Writing – Review & Editing, Resources, Project administration, Funding acquisition. TRJ: Conceptualization, Methodology, Resources, Writing – Review & Editing, Supervision, Project administration, Funding acquisition.

