## Supplemental Information for "Modelling exposure to aerosols from showers: implications for microbial risk assessment"

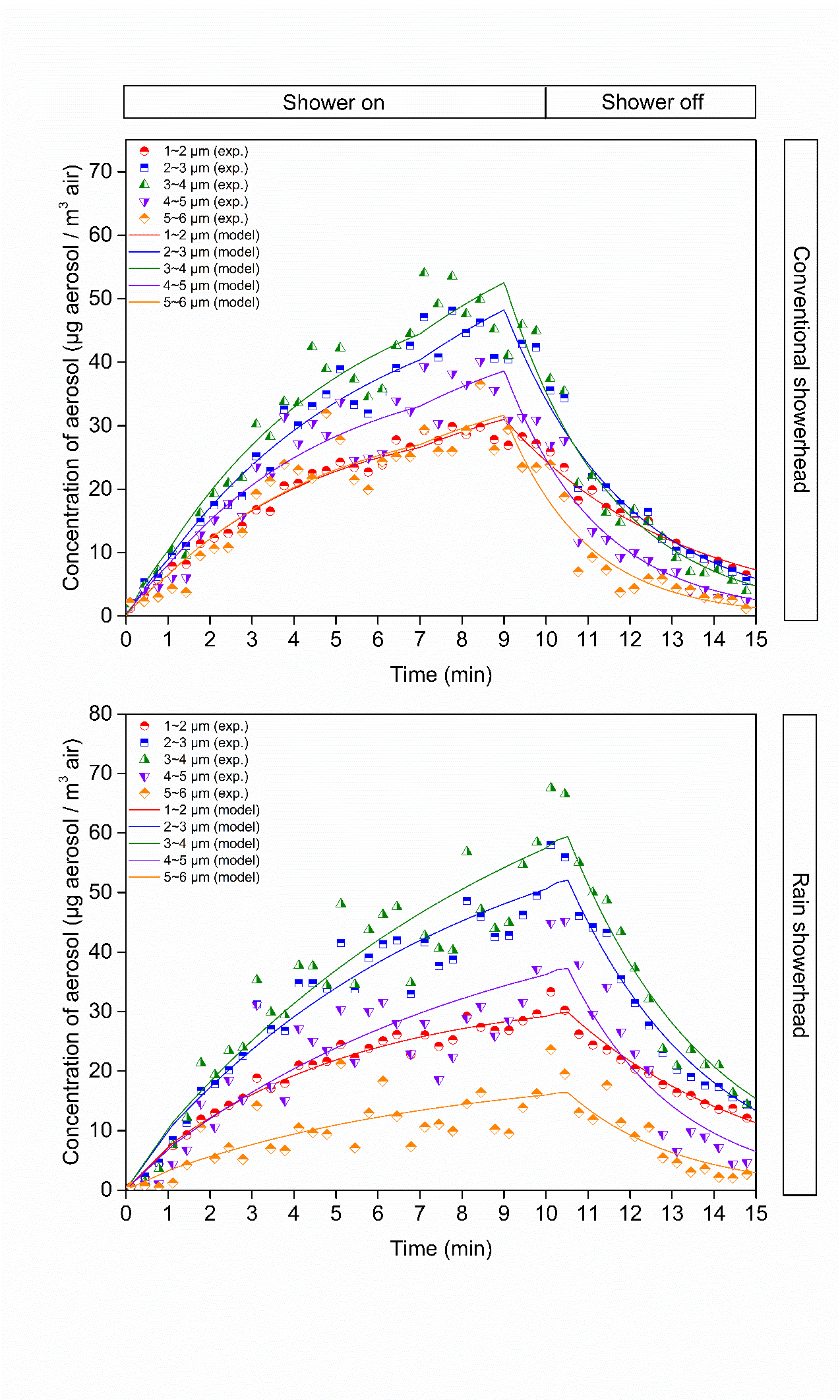

Figure S1. Concentration of aerosols of each size class for one of the triplicates at cold water temperature. The term exp refers to the experimentally collected data, and model refers to the model fit values.

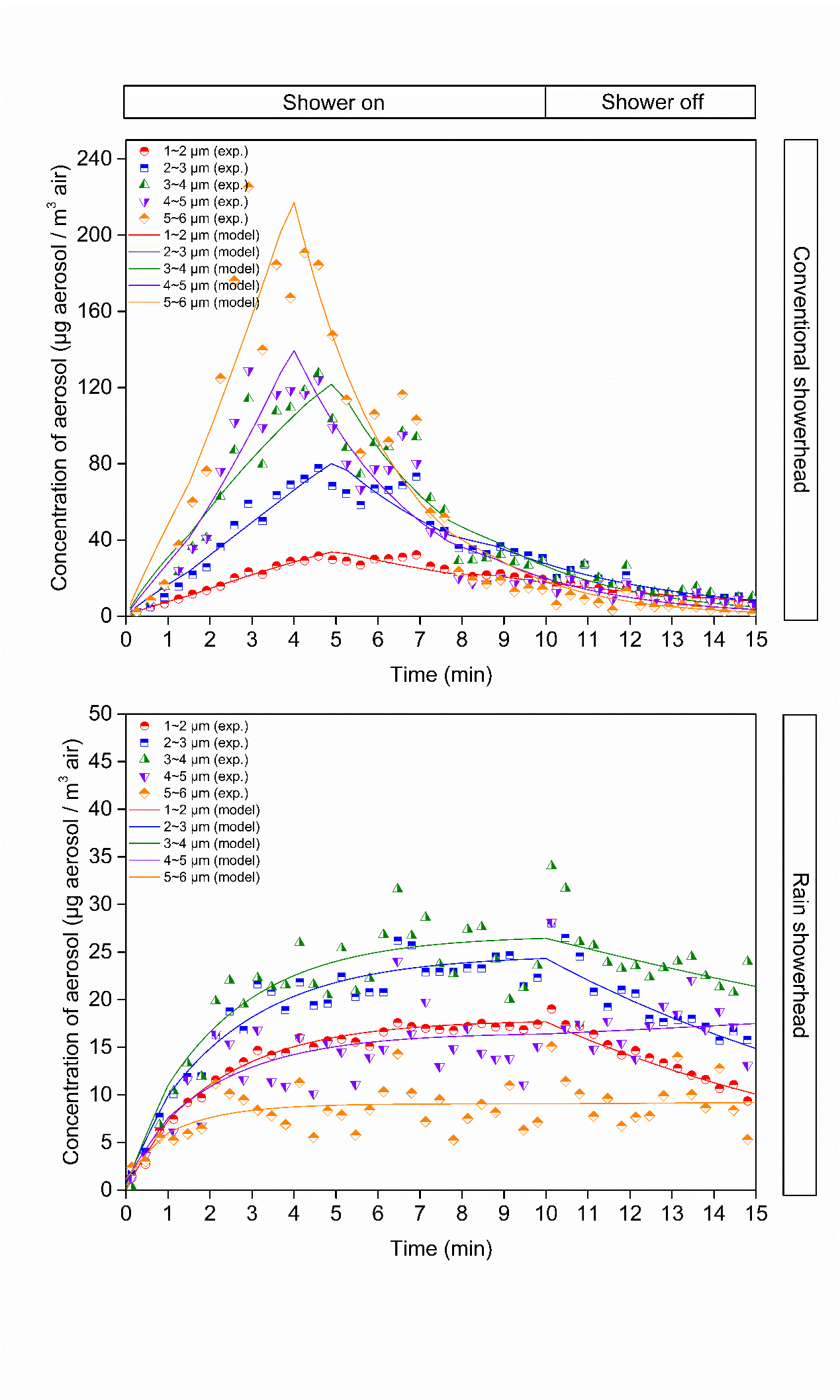

Figure S2. Concentration of aerosols of each size class for one of the triplicates at cold water temperature. The term exp refers to the experimentally collected data, and model refers to the model fit values.

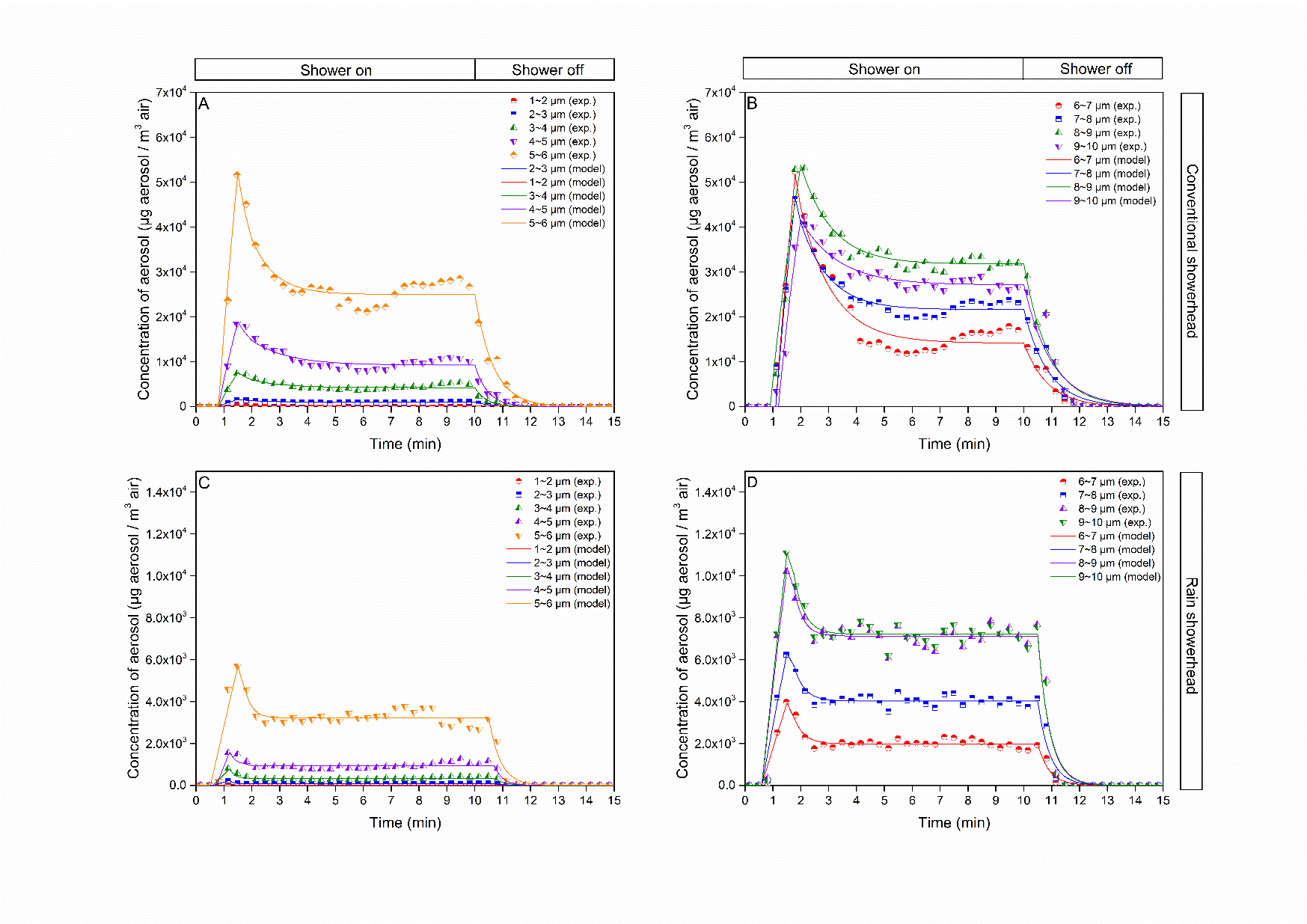

Figure S3. Concentration of aerosols of each size class for one of the triplicates at hot water temperature. The term exp refers to the experimentally collected data, and model refers to the model fit values.

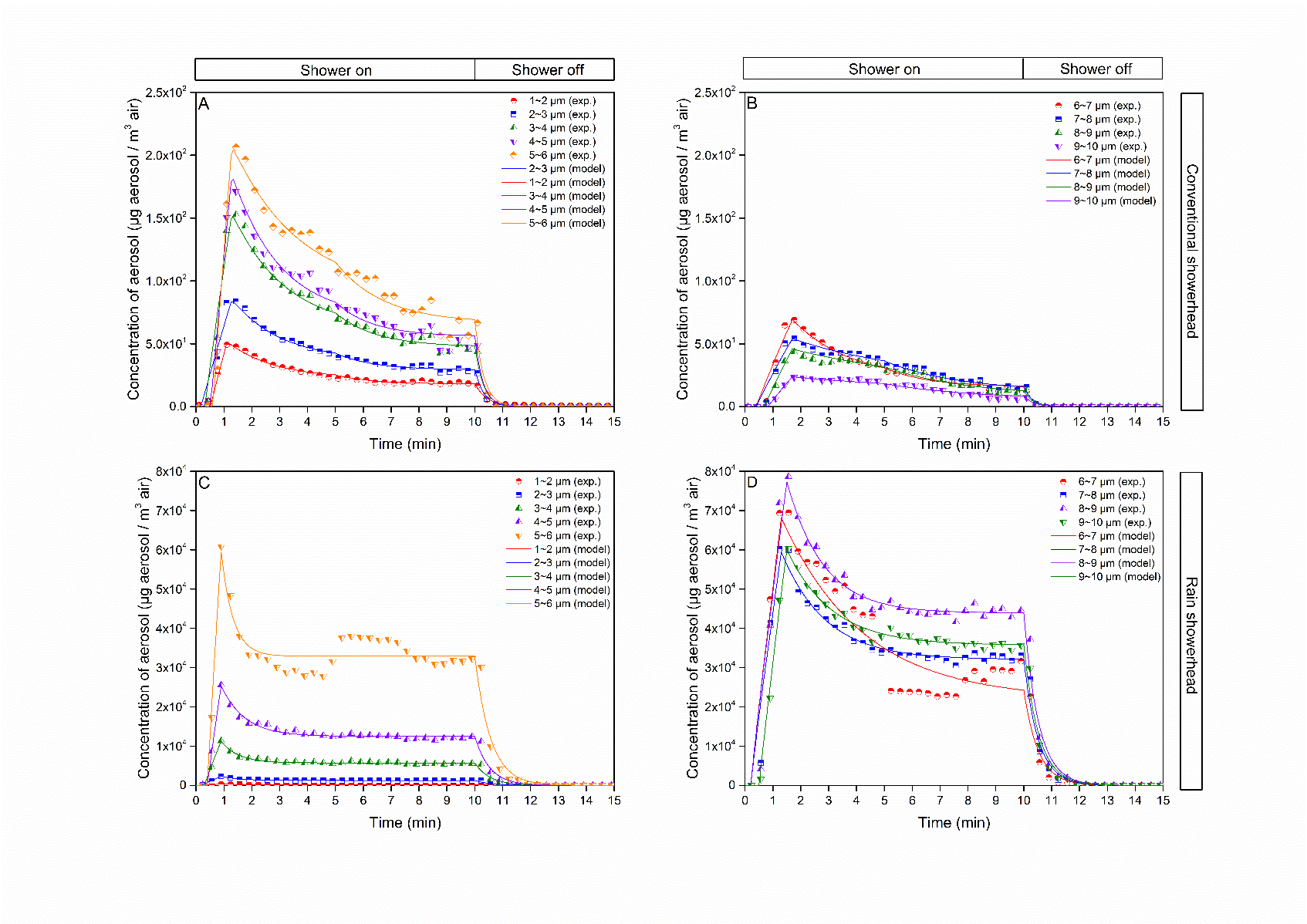

Figure S4. Concentration of aerosols of each size class for one of the triplicates at hot water temperature. The term exp refers to the experimentally collected data, and model refers to the model fit values.

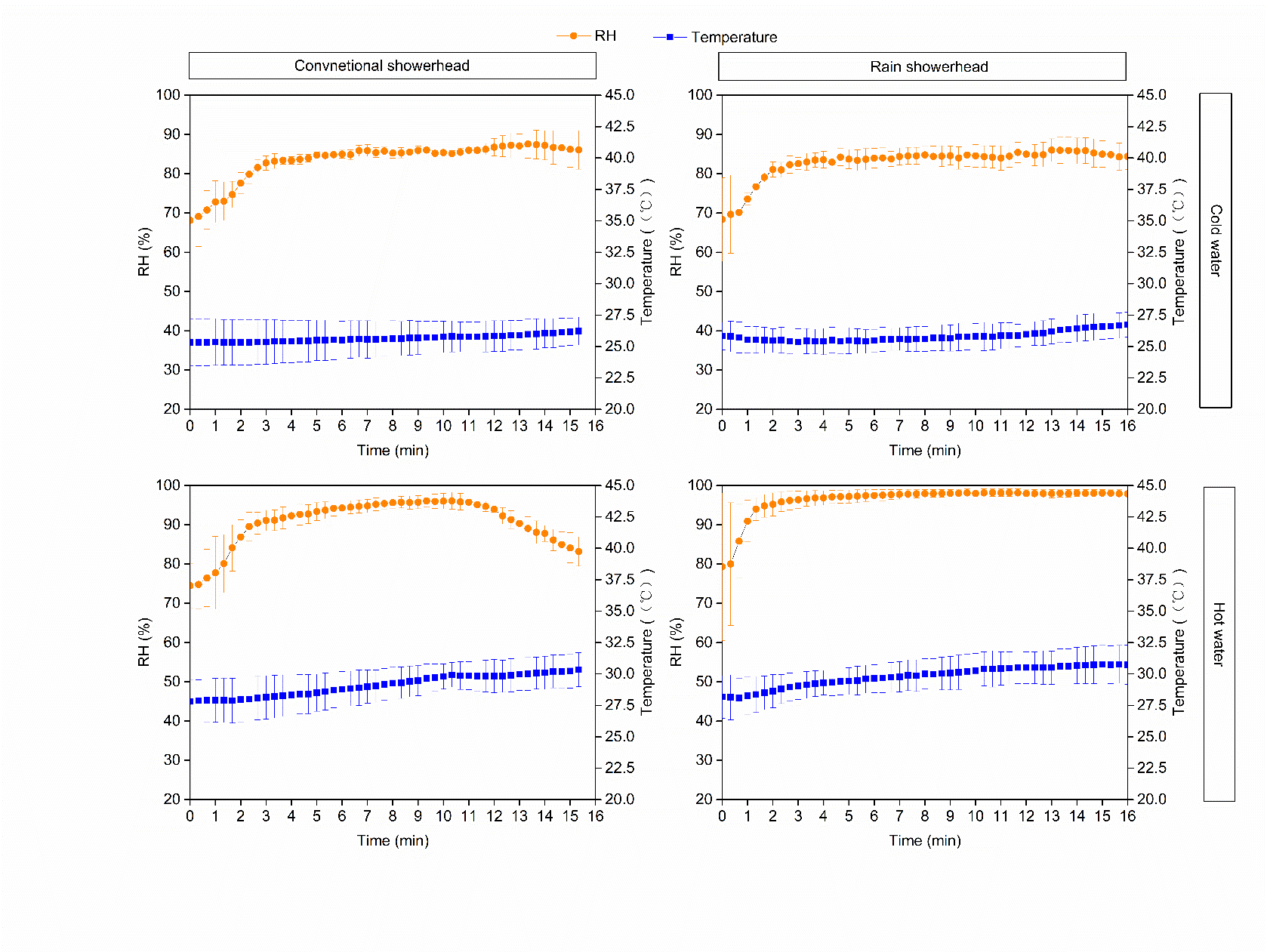

Figure S5. Measured air RH and temperature during showers. Error bars represent standard deviations. RH refers to relative humidity.

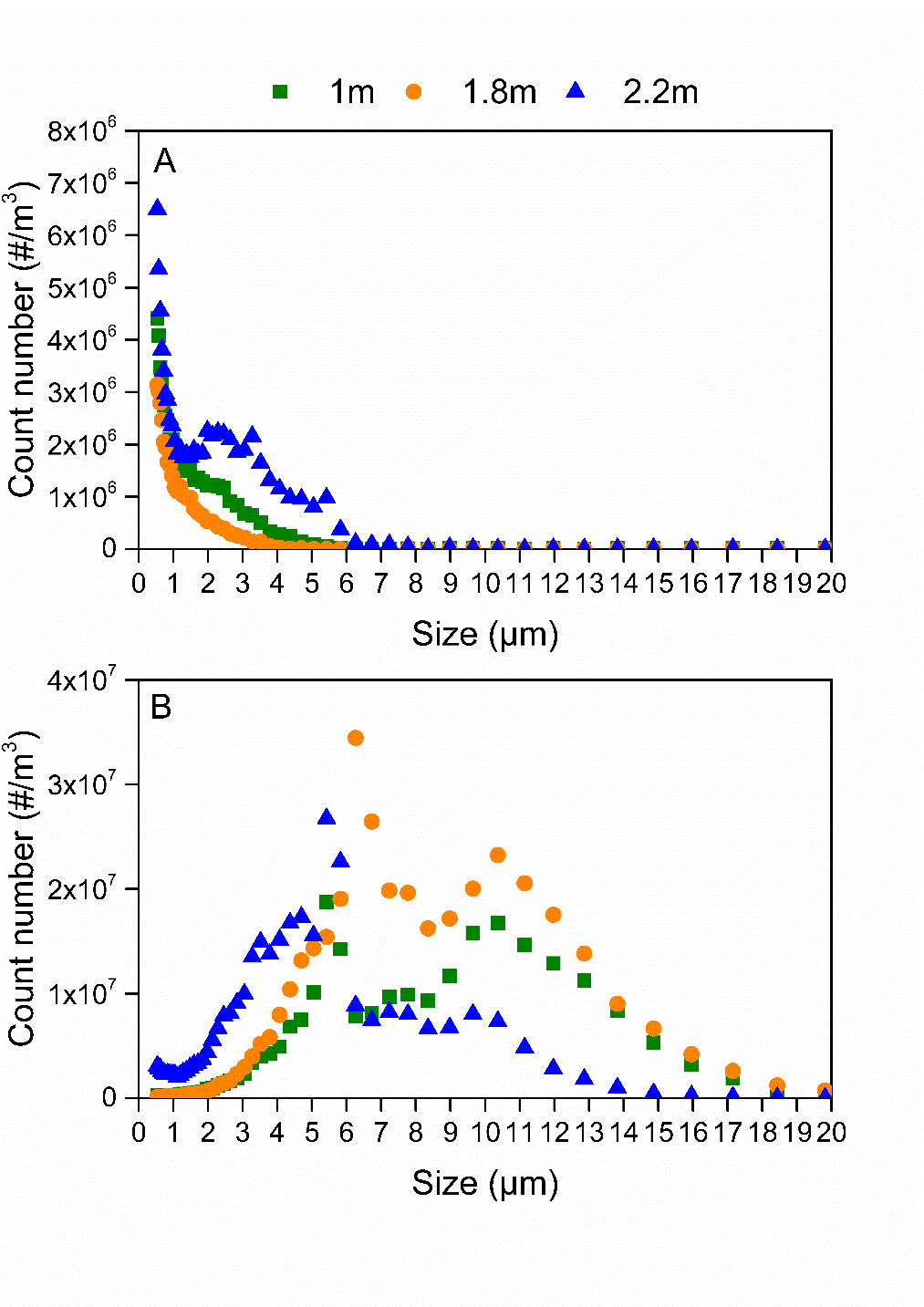

Figure S6. Steady state aerosol concentrations sampled at different locations within shower stall at cold water temperature (A) and hot water temperature (B)

Table S1. Exposure model parameters

| Parameter | Symbol | Distribution | Value | Unit | Source |
| --- | --- | --- | --- | --- | --- |
| Aerosol generation rate | G_i_ |  |  |  |  |
| Cold water |  |  |  |  |  |
| Conventional showerhead |  |  |  |  |  |
| 1-2 μm |  | Uniform | min=2.0e-2, max=2.3e-2 | mg/min | Model fitted |
| 2-3 μm |  | Uniform | min=2.7e-2, max=4.4e-2 | mg/min | Model fitted |
| 3-4 μm |  | Uniform | min=3.1e-2, max=8.7e-2 | mg/min | Model fitted |
| 4-5 μm |  | Uniform | min=2.5e-2, max=6.8e-2 | mg/min | Model fitted |
| 5-6 μm |  | Uniform | min=2.0e-2, max=1.2e-1 | mg/min | Model fitted |
| Rain showerhead |  |  |  |  | Model fitted |
| 1-2 μm |  | Uniform | min=1.7e-2, max=2.2e-2 | mg/min | Model fitted |
| 2-3 μm |  | Uniform | min=2.2e-2, max=3.0e-2 | mg/min | Model fitted |
| 3-4 μm |  | Uniform | min=2.3e-2, max=3.5e-2 | mg/min | Model fitted |
| 4-5 μm |  | Uniform | min=1.5e-2, max=2.7e-2 | mg/min | Model fitted |
| 5-6 μm |  | Uniform | min=7e-3, max=3.1e-2 | mg/min | Model fitted |
| Hot water |  |  |  |  |  |
| Conventional showerhead |  |  |  |  |  |
| The first minute |  |  |  |  |  |
| 1-2 μm |  | Uniform | min=2.2e-1, max=1.0 | mg/min | Model fitted |
| 2-3 μm |  | Uniform | min=2.0e-1, max=6.6 | mg/min | Model fitted |
| 3-4 μm |  | Uniform | min=4.1e-1, max=3.1e1 | mg/min | Model fitted |
| 4-5 μm |  | Uniform | min=6.2e-1, max=7.6e1 | mg/min | Model fitted |
| 5-6 μm |  | Uniform | min=7.9e-1, max=2.1e2 | mg/min | Model fitted |
| 6-7 μm |  | Uniform | min=1.4e-1, max=2.1e2 | mg/min | Model fitted |
| 7-8 μm |  | Uniform | min=1.1e-1, max=1.9e2 | mg/min | Model fitted |
| 8-9 μm |  | Uniform | min=1.2e-1, max=1.5e2 | mg/min | Model fitted |
| 9-10 μm |  | Uniform | min=7.2e-2, max=1.7e2 | mg/min | Model fitted |
| After the first minute |  |  |  |  |  |
| 1-2 μm |  | Uniform | min=3.4e-2, max=5.6e-1 | mg/min | Model fitted |
| 2-3 μm |  | Uniform | min=5.3e-2, max=2.3 | mg/min | Model fitted |
| 3-4 μm |  | Uniform | min=8.0e-2, max=9.4 | mg/min | Model fitted |
| 4-5 μm |  | Uniform | min=1.0e-1, max=1.8e1 | mg/min | Model fitted |
| 5-6 μm |  | Uniform | min=9.5e-2, max=7.5e1 | mg/min | Model fitted |
| 6-7 μm |  | Uniform | min=2.4e-2, max=30.3 | mg/min | Model fitted |
| 7-8 μm |  | Uniform | min=1.1, max=5.3e1 | mg/min | Model fitted |
| 8-9 μm |  | Uniform | min=7.2e-3, max=7.2e1 | mg/min | Model fitted |
| 9-10 μm |  | Uniform | min=4.6e-3, max=5.0e1 | mg/min | Model fitted |
| Rain showerhead |  |  |  |  |  |
| The first minute |  |  |  |  |  |
| 1-2 μm |  | Uniform | min=1.7e-1, max=1.32 | mg/min | Model fitted |
| 2-3 μm |  | Uniform | min=9.5e-1, max=9.1 | mg/min | Model fitted |
| 3-4 μm |  | Uniform | min=3.2, max=5.0e1 | mg/min | Model fitted |
| 4-5 μm |  | Uniform | min=8.2, max=1.4e2 | mg/min | Model fitted |
| 5-6 μm |  | Uniform | min=14.6, max=3.2e2 | mg/min | Model fitted |
| 6-7 μm |  | Uniform | min=11.0, max=1.7e2 | mg/min | Model fitted |
| 7-8 μm |  | Uniform | min=17.7, max=1.5e2 | mg/min | Model fitted |
| 8-9 μm |  | Uniform | min=29.2, max=1.7e2 | mg/min | Model fitted |
| 9-10 μm |  | Uniform | min=31.4, max=1.7e2 | mg/min | Model fitted |
| After the first minute |  |  |  |  |  |
| 1-2 μm |  | Uniform | min=2.6e-2, max=5.8e-1 | mg/min | Model fitted |
| 2-3 μm |  | Uniform | min=1.5e-1, max=4.7 | mg/min | Model fitted |
| 3-4 μm |  | Uniform | min=8.2e-1, max=2.3e1 | mg/min | Model fitted |
| 4-5 μm |  | Uniform | min=3.0, max=3.6e1 | mg/min | Model fitted |
| 5-6 μm |  | Uniform | min=9.8, max=1.9e2 | mg/min | Model fitted |
| 6-7 μm |  | Uniform | min=8.9, max=1.9e-1 | mg/min | Model fitted |
| 7-8 μm |  | Uniform | min=1.7e1, max=5.0e1 | mg/min | Model fitted |
| 8-9 μm |  | Uniform | min=2.1e1, max=8.3e1 | mg/min | Model fitted |
| 9-10 μm |  | Uniform | min=2.4e1, max=5.5e1 | mg/min | Model fitted |
| Aerosol residual decay rate | λ_i_ |  |  |  |  |
| Cold water |  |  |  |  |  |
| Conventional showerhead |  |  |  |  |  |
| During shower |  |  |  |  |  |
| 1-2 μm |  | Uniform | min=-3.4e-2, max=1.5e-1 | 1/min | Model fitted |
| 2-3 μm |  | Uniform | min=-9.0e-3, max=7.0e-3 | 1/min | Model fitted |
| 3-4 μm |  | Uniform | min=-2.3e-2, max=2.3e-2 | 1/min | Model fitted |
| 4-5 μm |  | Uniform | min=-2.7e-1, max=3.9e-2 | 1/min | Model fitted |
| 5-6 μm |  | Uniform | min=-2.1e-1, max=6.5e-2 | 1/min | Model fitted |
| After shower |  |  |  |  |  |
| 1-2 μm |  | Uniform | min=3.3e-2, max=4.5e-1 | 1/min | Model fitted |
| 2-3 μm |  | Uniform | min=1.2e-1, max=7.2e-1 | 1/min | Model fitted |
| 3-4 μm |  | Uniform | min=2.0e-1, max=9.1e-1 | 1/min | Model fitted |
| 4-5 μm |  | Uniform | min=2.1e-1, max=1.1 | 1/min | Model fitted |
| 5-6 μm |  | Uniform | min=2.9e-1, max=1.2 | 1/min | Model fitted |
| Rain showerhead |  |  |  |  |  |
| During shower |  |  |  |  |  |
| 1-2 μm |  | Uniform | min=-1.0e-1, max=1.7e-1 | 1/min | Model fitted |
| 2-3 μm |  | Uniform | min=-1.8e-1, max=1.3e-1 | 1/min | Model fitted |
| 3-4 μm |  | Uniform | min=-2.1e-1, max=1.6e-1 | 1/min | Model fitted |
| 4-5 μm |  | Uniform | min=-2.0e-1, max=2.8e-1 | 1/min | Model fitted |
| 5-6 μm |  | Uniform | min=-1.9e-1, max=1.1 | 1/min | Model fitted |
| After shower |  |  |  |  |  |
| 1-2 μm |  | Uniform | min=-5.3e-2, max=2.9e-1 | 1/min | Model fitted |
| 2-3 μm |  | Uniform | min=3.1e-2, max=3.4e-1 | 1/min | Model fitted |
| 3-4 μm |  | Uniform | min=2.4e-2, max=3.2e-1 | 1/min | Model fitted |
| 4-5 μm |  | Uniform | min=-3.1e-2, max=4.0e-1 | 1/min | Model fitted |
| 5-6 μm |  | Uniform | min=-3.0e-2, max=3.0e-1 | 1/min | Model fitted |
| Hot water |  |  |  |  |  |
| Conventional showerhead |  |  |  |  |  |
| During shower |  |  |  |  |  |
| 1-2 μm |  | Uniform | min=5.1e-1, max=1.2 | 1/min | Model fitted |
| 2-3 μm |  | Uniform | min=4.7e-1, max=8.8e-1 | 1/min | Model fitted |
| 3-4 μm |  | Uniform | min=4.7e-1, max=8.8e-1 | 1/min | Model fitted |
| 4-5 μm |  | Uniform | min=4.0e-1, max=6.3e-1 | 1/min | Model fitted |
| 5-6 μm |  | Uniform | min=4.7e-1, max=5.0e-1 | 1/min | Model fitted |
| 6-7 μm |  | Uniform | min=1.7e-1, max=9.4e-1 | 1/min | Model fitted |
| 7-8 μm |  | Uniform | min=3.4e-1, max=8.0e-1 | 1/min | Model fitted |
| 8-9 μm |  | Uniform | min=7.4e-2, max=6.9e-1 | 1/min | Model fitted |
| 9-10 μm |  | Uniform | min=-8.3e-3, max=4.2e-1 | 1/min | Model fitted |
| After shower |  |  |  |  |  |
| 1-2 μm |  | Uniform | min=1.2, max=2.0 | 1/min | Model fitted |
| 2-3 μm |  | Uniform | min=1.2, max=2.8 | 1/min | Model fitted |
| 3-4 μm |  | Uniform | min=1.7, max=3.1 | 1/min | Model fitted |
| 4-5 μm |  | Uniform | min=1.9, max=3.3 | 1/min | Model fitted |
| 5-6 μm |  | Uniform | min=1.3, max=3.0 | 1/min | Model fitted |
| 6-7 μm |  | Uniform | min=8.9e-1, max=3.1 | 1/min | Model fitted |
| 7-8 μm |  | Uniform | min=8.6e-1, max=2.9 | 1/min | Model fitted |
| 8-9 μm |  | Uniform | min=8.0e-1, max=2.8 | 1/min | Model fitted |
| 9-10 μm |  | Uniform | min=6.8e-1, max=2.5 | 1/min | Model fitted |
| Rain showerhead |  |  |  |  |  |
| During shower |  |  |  |  |  |
| 1-2 μm |  | Uniform | min=1.2, max=5.3 | 1/min | Model fitted |
| 2-3 μm |  | Uniform | min=1.1, max=8.3 | 1/min | Model fitted |
| 3-4 μm |  | Uniform | min=1.1, max=6.6 | 1/min | Model fitted |
| 4-5 μm |  | Uniform | min=1.0, max=5.3 | 1/min | Model fitted |
| 5-6 μm |  | Uniform | min=8.9e-1, max=3.2 | 1/min | Model fitted |
| 6-7 μm |  | Uniform | min=1.6e-1, max=2.3 | 1/min | Model fitted |
| 7-8 μm |  | Uniform | min=4.3e-1, max=2.2 | 1/min | Model fitted |
| 8-9 μm |  | Uniform | min=5.5e-1, max=2.6 | 1/min | Model fitted |
| 9-10 μm |  | Uniform | min=3.9e-1, max=2.2 | 1/min | Model fitted |
| After shower |  |  |  |  |  |
| 1-2 μm |  | Uniform | min=2.2e-1, max=1.1 | 1/min | Model fitted |
| 2-3 μm |  | Uniform | min=1.2, max=1.6 | 1/min | Model fitted |
| 3-4 μm |  | Uniform | min=1.6, max=3.9 | 1/min | Model fitted |
| 4-5 μm |  | Uniform | min=1.6, max=7.0 | 1/min | Model fitted |
| 5-6 μm |  | Uniform | min=1.5, max=1.3e1 | 1/min | Model fitted |
| 6-7 μm |  | Uniform | min=1.6, max=1.3e1 | 1/min | Model fitted |
| 7-8 μm |  | Uniform | min=1.6, max=1.3e1 | 1/min | Model fitted |
| 8-9 μm |  | Uniform | min=1.6, max=1.3e1 | 1/min | Model fitted |
| 9-10 μm |  | Uniform | min=1.6, max=1.3e1 | 1/min | Model fitted |
| Volume of shower stall | V_m_ | Point | 2.5 | m^3^ | Measured |
| Ventilation rate | Q |  |  |  |  |
| Cold water conventional showerhead |  |  |  |  |  |
| Before shower |  | Uniform | min=8.5e-1, max=9.6e- | m^3^/min | Measured |
| During shower |  | Uniform | min=2.4e-1, max=5.5e-1 | m^3^/min | Measured |
| After shower |  | Uniform | min=1.1e-1, max=5.5e-1 | m^3^/min | Measured |
| Cold water rain showerhead |  |  |  |  |  |
| Before shower |  | Uniform | min=8.8e-2, max=5.5e-1 | m^3^/min | Measured |
| During shower |  | Uniform | min=6.8e-1, max=1.5 | m^3^/min | Measured |
| After shower |  | Uniform | min=0.0, max=6.6e-1 | m^3^/min | Measured |
| Hot water conventional showerhead |  |  |  |  |  |
| Before shower |  | Uniform | min=0.0, max=9.8e-1 | m^3^/min | Measured |
| During shower |  | Uniform | min=2.5e-1, max=6.3e-1 | m^3^/min | Measured |
| After shower |  | Uniform | min=6.2e-1, max=1.2 | m^3^/min | Measured |
| Hot water rain showerhead |  |  |  |  |  |
| Before shower |  | Uniform | min=0.0, max=4.4e-1 | m^3^/min | Measured |
| During shower |  | Uniform | min=3.7e-1, max=1.1 | m^3^/min | Measured |
| After shower |  | Uniform | min=3.4e-2, max=1.1 | m^3^/min | Measured |
| Inhalation rate | Br | Uniform | min=1.3e-2, max=1.7e-2 | m^3^/min | (Epa, 2011) |
| Deposition efficiency | D_i_ |  |  |  | (Heyder et al., 1986) |
| 1-2 μm |  | Uniform | min=2.3e-1, max=5.3e-1 | Unitless |  |
| 2-3 μm |  | Uniform | min=3.6e-1, max=6.2e-1 | Unitless |  |
| 3-4 μm |  | Uniform | min=2.9e-1, max=6.2e-1 | Unitless |  |
| 4-5 μm |  | Uniform | min=1.9e-1, max=6.1e-1 | Unitless |  |
| 5-6 μm |  | Uniform | min=1.0e-1, max=5.2e-1 | Unitless |  |
| 6-7 μm |  | Uniform | min=3.0e-2, max=4.0e-1 | Unitless |  |
| 7-8 μm |  | Uniform | min=3.0e-2, max=2.9e-1 | Unitless |  |
| 8-9 μm |  | Uniform | min=1.0e-2, max=1.9e-1 | Unitless |  |
| 9-10 μm |  | Uniform | min=1.0e-2, max=1.2e-1 | Unitless |  |
